# Semantic computational analysis of anticoagulation use in atrial fibrillation from real world data

**DOI:** 10.1101/19011643

**Authors:** Daniel M. Bean, James Teo, Honghan Wu, Ricardo Oliveira, Raj Patel, Rebecca Bendayan, Ajay M. Shah, Richard J. B. Dobson, Paul A. Scott

## Abstract

Atrial fibrillation (AF) is the most common arrhythmia and significantly increases stroke risk. This risk is effectively managed by oral anticoagulation. Recent studies using national registry data indicate increased use of anticoagulation resulting from changes in guidelines and the availability of newer drugs.

The aim of this study is to develop and validate an open source risk scoring pipeline for free-text electronic health record data using natural language processing.

AF patients discharged from 1^st^ January 2011 to 1^st^ October 2017 were identified from discharge summaries (N=10,030, 64.6% male, average age 75.3 ± 12.3 years). A natural language processing pipeline was developed to identify risk factors in clinical text and calculate risk for ischaemic stroke (CHA_2_DS_2_-VASc) and bleeding (HAS-BLED). Scores were validated vs two independent experts for 40 patients.

Automatic risk scores were in strong agreement with the two independent experts for CHA_2_DS_2_-VASc (average kappa 0.78 vs experts, compared to 0.85 between experts). Agreement was lower for HAS-BLED (average kappa 0.54 vs experts, compared to 0.74 between experts).

In high-risk patients (CHA_2_DS_2_-VASc ≥2) OAC use has increased significantly over the last 7 years, driven by the availability of DOACs and the transitioning of patients from AP medication alone to OAC. Factors independently associated with OAC use included components of the CHA_2_DS_2_-VASc and HAS-BLED scores as well as discharging specialty and frailty. OAC use was highest in patients discharged under cardiology (69%).

Electronic health record text can be used for automatic calculation of clinical risk scores at scale. Open source tools are available today for this task but require further validation. Analysis of routinely-collected EHR data can replicate findings from large-scale curated registries.

## Introduction

Atrial fibrillation (AF) affects 2% of the UK population and significantly increases stroke risk.[1] Although this risk can be substantially reduced by oral anticoagulants (OAC), warfarin has historically been underused in AF. Over the last decade the antithrombotic landscape has changed significantly with: (1) the introduction of direct oral anticoagulants (DOACs), and (2) the updated UK NICE 2014 AF guidelines[2] which introduced the CHA_2_DS_2_-VASc[3] and HAS-BLED[4] risk calculators and removed endorsement of the use of antiplatelet agents for stroke prevention. A number of large-scale observational studies have found that rates of OAC use have significantly increased since the introduction of DOACs.[5–8] However, these previous analyses have used structured data, which do not capture the full clinical narrative, and many studies have used registry data which can be costly and time-consuming to collect and may not always accurately reflect real-world practice.

An alternative approach to observational research is the use of Electronic Health Record (EHRs) data generated as part of routine clinical care.[9] Modern EHRs contain a combination of structured (e.g. age, sex) and unstructured (e.g. free text, image) data. Whilst free text is information-dense to a human reader, to be useful for computational analysis it requires conversion to a structured format. Performing this process manually is very labour-intensive. However, given the enormous volume of clinical data contained solely in written notes[10], extracting this information is critical to realizing the full potential of EHRs.

Natural language processing (NLP) uses computer algorithms to identify key elements in everyday language and extract meaning from spoken or written language. NLP can be used to convert unstructured text found in EHRs to structured data. This should allow rapid, low-cost and automated analysis of medical text, including the generation of observational data for research purposes.

In this study we develop an NLP pipeline to calculate clinical risk scores from free text. We build upon our existing data pooling, harmonization and information retrieval tool (CogStack[11,12]), together with a semantic NLP tool for information extraction (SemEHR[13,14]). Previous studies have found it is possible to accurately predict CHA_2_DS_2_-VASc using EHR text.[15–17] We build on this work to develop a flexible open source pipeline and calculate additional risk scores. Our specific objectives are to:

a. Develop and validate an NLP risk scoring pipeline.
b. Explore trends in antithrombotic medication use for AF including the impact of the availability of DOACs and changes in NICE 2014 guidelines.
c. Quantify the association between antithrombotic medication use and relevant clinical patient-level variables.

## Methods

### Data, materials and code

A subset of the dataset limited to anonymisable information (e.g. only UMLS codes and demographics) is available on request to researchers with suitable training in information governance and human confidentiality protocols; contact. All code for calculating risk scores is open-source in GitHub at https://github.com/CogStack/risk-score-builder. Source text from patient records used in the study will not be available due to inability to fully anonymise up to the Information Commissioner Office (ICO) standards. Risk factor-level data is available as S3 Table.

### Ethical approval

This study was performed on anonymised data as a clinical audit for service evaluation. The project was reviewed by the King’s College Hospital Information Governance committee chaired by the Caldicott Guardian Professor Alastair Baker (the Caldicott Guardian is the statutory individual responsible for protecting the confidentiality of health and care information in a UK healthcare organisation) and approval was granted in November 2018 with continued oversight. The legal basis of secondary use was analysis for service evaluation, operational performance and clinical audit.

### Cohort selection

We used an open-source retrieval system for unstructured clinical data (CogStack)[11,12] to define a cohort of patients aged ≥ 18 with AF admitted to KCH between 01-01-2011 and 01-10-2017. We searched discharge summaries for adult inpatients discharged alive containing the exact keywords “AF”, “PAF”, “AFib” or “Atrial Fibrillation”. Although the risk of stroke and OAC indications in atrial flutter are similar to AF, in clinical practice in the UK many patients with isolated typical flutter undergo flutter ablation after which there is significant variation in practice in terms of long-term OAC prescription. For this reason we decided not to include patients with flutter. Patients with missing data such as gender or discharge ward were excluded (N=397). We also excluded patients discharged directly from the emergency department, day units or the clinical decision unit, as these did not constitute an inpatient admission and did not generate the discharge summaries we used to identify discharge medication and diagnosis of AF.

We further refined our cohort using an NLP pipeline SemEHR[13,14] which generates semantic annotation and can detect negation, temporality (current, historic) and experiencer. We excluded patients for which the NLP pipeline detected negation, a hypothetical mention or another experiencer (the mention refers to another individual who is not the patient e.g. family history) for AF.

We defined a new diagnosis of AF as the first mention of AF in a patient with at least one previous visit and no earlier record of AF or prescription of antithrombotic medication.

### CHA2DS2-VASc and HAS-BLED risk score calculation

We used the SemEHR NLP pipeline to annotate clinical documents with Unified Medical Language System (UMLS) concepts.[18] To calculate CHA_2_DS_2_-VASc and HAS-BLED risk scores, we manually mapped each phenotypic component of the score (e.g. stroke) to the closest general term in the Human Phenotype Ontology (HPO)[19] and automatically included all descendent terms in the ontology. All HPO concepts were then mapped automatically to UMLS. Medications were manually mapped to UMLS concepts directly (as they are not present in HPO), and the first child terms are included automatically using UMLS concept relationships. The only factor not included was a labile International Normalised Ratio (INR) in the HAS-BLED score, which is not in HPO and is ambiguous in UMLS, and which is not reliably recorded in the dataset.

The result is a mapping of each score component to a list of UMLS concepts, which was manually refined based on manual review of a random sample of 205 patients by a single annotator. The final mapping is available as S1 Table. For each component we then identified matching annotations in medical records using the NLP pipeline and awarded points as defined for each score.

For patients with multiple admissions (and the possibility of change in risk scores over time) we used the most recent admission to calculate risk scores.

### Antithrombotic Drug Prescription

Antithrombotic prescriptions of OACs (apixaban, rivaroxaban, dabigatran, edoxaban, warfarin) and antiplatelets (AP; aspirin, clopidogrel, dipyridamole, ticagrelor, prasugrel) were extracted from free text discharge summaries. This was performed using a custom NLP pipeline written in Python and specifically adapted to the KCH record structure. Drug mentions are identified by fuzzy matching and any detected mentions are tested for negation using regular expressions. The open source code is available at https://github.com/CogStack/OAC-NLP.

### Hospital Frailty Risk Score (HFRS) Calculation

We calculated the Hospital Frailty Risk Score (HFRS) proposed by Gilbert *et al*. [20] which uses ICD-10 diagnostic codes to identify a group of patients at higher risk of adverse outcomes. We mapped these ICD-10 codes to UMLS concept unique identifiers (CUI) using bio-ontology.[21] We used SemEHR to detect all UMLS concepts in free text and calculate the total frailty risk as the sum of concept weights as defined by Gilbert *et al*..[20]

### Validation of AF diagnosis, Antithrombotic drug prescription and NLP risk scores

The diagnosis of AF and antithrombotic drug prescriptions were manually validated on a random sample of 300 discharge summaries (AF diagnosis) or 200 discharge summaries (prescription) taken from our cohort. Performance was measured by calculating the precision, recall and F1 score.

CHA_2_DS_2_-VASc and HAS-BLED risk scores were validated for a sample of 40 patients selected at random after stratification by gender and age (this sample does not overlap with the initial sample used to refine the automated scoring). Each patient was manually scored for all components of CHA_2_DS_2_-VASc and HAS-BLED by two independent expert clinicians according to agreed criteria (see S1 Table). Inter-annotator agreement for the final scores was calculated using a weighted Cohen’s kappa. Given the high-dimensional complexity of the HFRS, we did not attempt to validate it and instead compared the score distribution to the original findings of Gilbert *et al*..[20]

### Statistical analysis

Categorical variables are expressed as percentages and compared using a chi-squared test. Normally distributed continuous variables are expressed as mean+/-standard deviation and compared using Student *t* test. Skewed continuous variables (length of stay, number of visits, HFRS) are expressed as median (minimum-maximum) and compared using a Kruskal-Wallis H-test. Statistical analyses were performed using the StatsModels and scipy libraries in Python. In all analyses a *P*<0.05 was considered significant.

We evaluated temporal trends in the rates of prescription of antithrombotic drugs for patients at high stroke risk (CHA_2_DS_2_-VASc ≥ 2) using linear regression with quarterly data, retaining the last visit per quarter for each patient.

The association of individual risk score (CHA_2_DS_2_-VASc and HAS-BLED) components and other clinical variables with antithrombotic prescription were evaluated in univariate and multivariate analyses. Factors with a significant association (P<0.05) in univariate analysis were entered into multivariate models. These associations were estimated using odds ratios from logistic regression. Uncontrolled hypertension and concomitant alcohol abuse were not included in the models as there were too few positive cases in our validation data. Concomitant drugs increasing bleeding risk were also excluded as this includes antiplatelets which could be prescribed for anticoagulation.

## Results

### Cohort identification

We identified 11,260 adult patients admitted to KCH with a diagnosis of AF. After excluding 1,230 patients (Fig 1) we were left with a final cohort of 10,030 patients admitted 17,387 times during the prescribing study period and 151,174 times in total (Table 1).

**Table 1.**
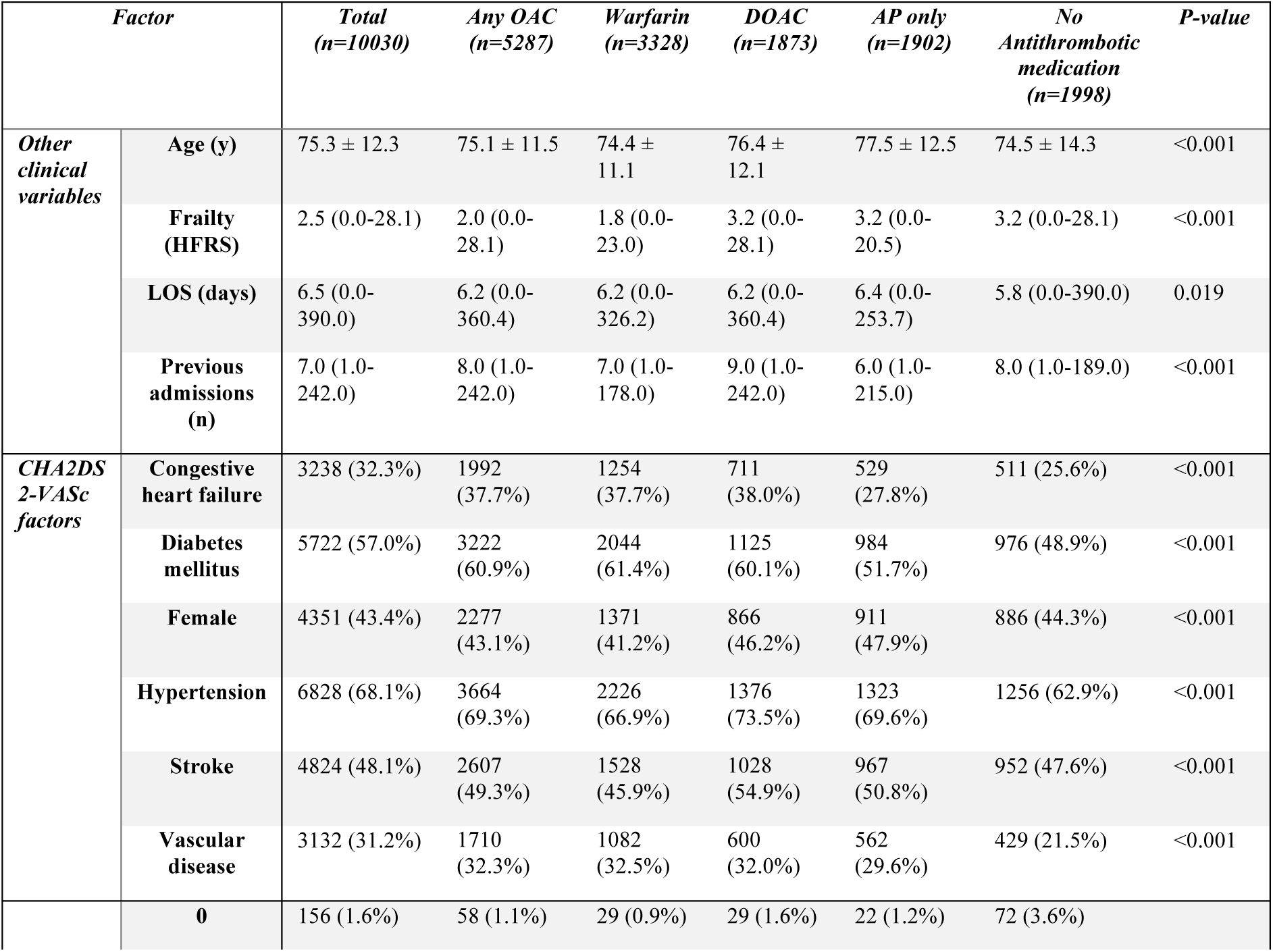
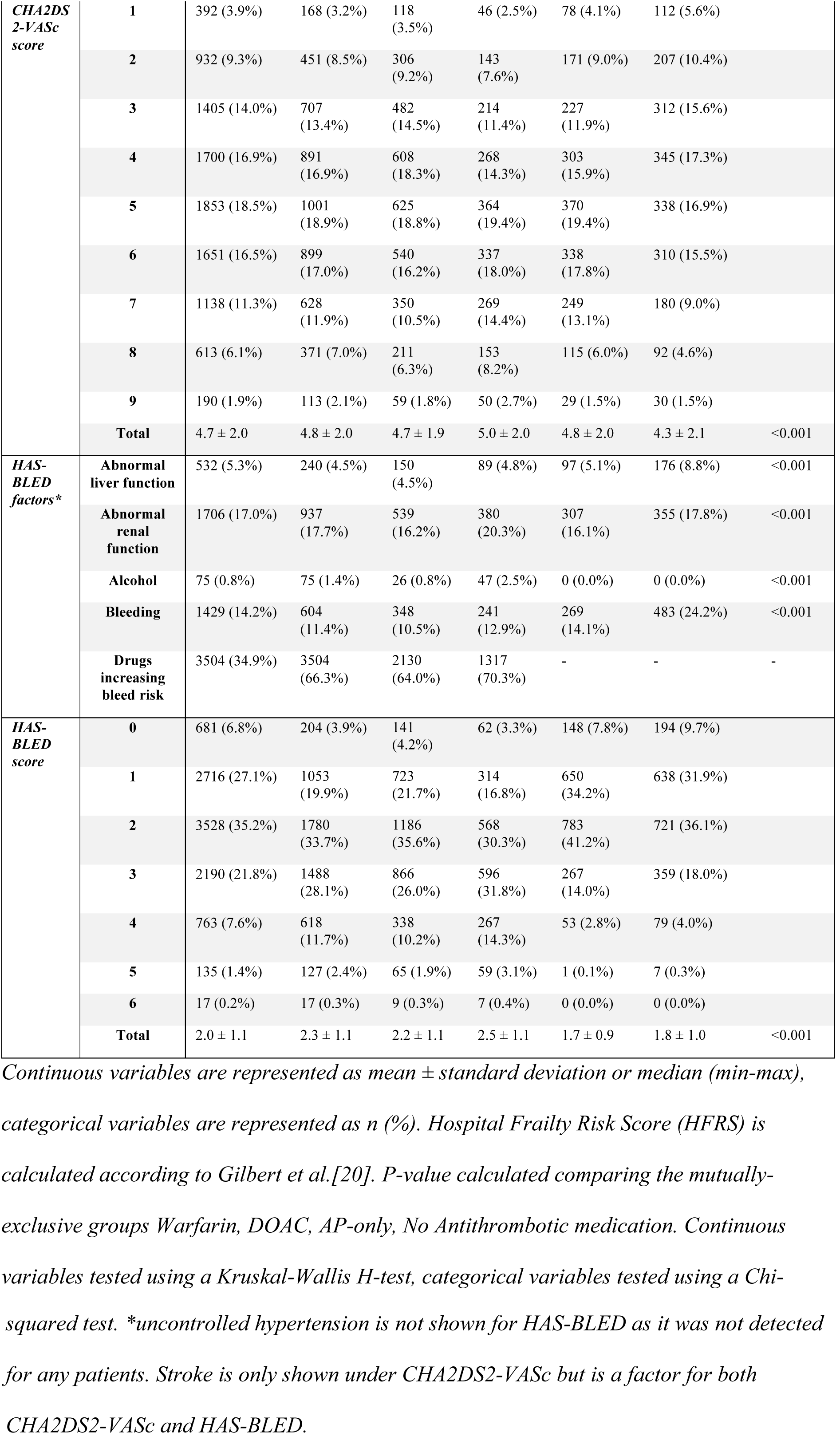
Baseline characteristics of study cohort.

**Fig 1.**
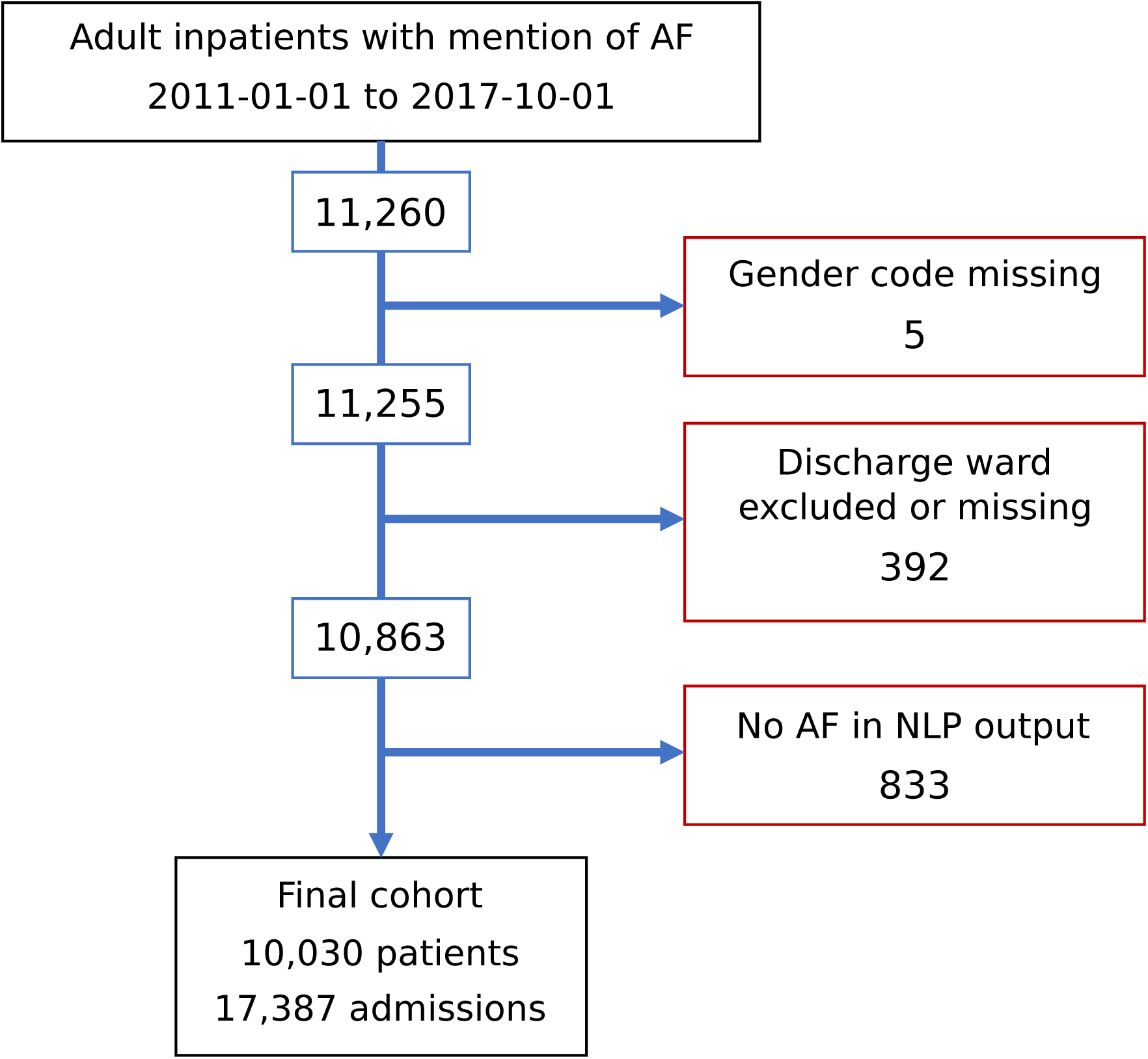
Derivation of the study cohort. AF = Atrial fibrillation, NLP = natural language processing.

### Validation of AF diagnosis, Antithrombotic drug prescription and NLP risk scores

A diagnosis of AF was confirmed in 96% of 300 cases reviewed. Of these, 200 cases were manually coded for prescription of any of 10 antithrombotic medications. Five drugs with <5 positive examples in the validation sample were excluded (edoxaban, dipyridamole, prasugrel, dabigatran, ticagrelor) due to the small sample size. The pipeline achieved perfect precision and recall for these excluded drugs but the sample size was too small to be meaningful. The average performance over the remaining 5 drugs was 95% precision at 97% recall (Table 2).

**Table 2.** Performance of the drug NLP pipeline in manual validation.

| Drug | Accuracy | Precision | Recall | F1 | P | FN | FP | TN | TP |
| --- | --- | --- | --- | --- | --- | --- | --- | --- | --- |
| Warfarin | 0.94 | 0.87 | 0.97 | 0.92 | 69 | 2 | 10 | 121 | 67 |
| Aspirin | 0.96 | 0.90 | 0.98 | 0.94 | 62 | 1 | 7 | 131 | 61 |
| Rivaroxaban | 1.00 | 1.00 | 0.95 | 0.98 | 22 | 1 | 0 | 178 | 21 |
| Clopidogrel | 1.00 | 1.00 | 0.94 | 0.97 | 17 | 1 | 0 | 183 | 16 |
| Apixaban | 1.00 | 1.00 | 1.00 | 1.00 | 13 | 0 | 0 | 187 | 13 |
| Average | 0.98 | 0.95 | 0.97 | 0.96 |  |  |  |  |  |

Discharge summaries were selected at random (n=200) and manually annotated for the prescription of the 10 drugs detected by the pipeline. Performance for the 5 drugs with > 10 positive examples in manual annotation is shown. P = total positive examples in manual annotation, FN = false negative, FP = false positive, TN = true negative, TP = true positive.

The performance of the automatic NLP scoring procedure was evaluated in 40 patients. Overall the agreement between two human expert raters and the algorithm for CHA_2_DS_2_-VASc was high for all pairs, and only slightly higher for the two human raters than for the algorithm vs. either expert. HAS-BLED agreement however was lower for all comparisons (Table 3 and S2 Table). Total scores and risk factor-level variables are available as S3 Table.

**Table 3.** Inter-rater agreement statistics for CHA_2_DS_2_-VASc and HAS-BLED risk scores.

| Score | Rater 1 | Rater 2 | Kappa (95% CI) |
| --- | --- | --- | --- |
| CHA<sub>2</sub>DS<sub>2</sub>-VASc | Algorithm | Expert A | 0.76 (0.65-0.86) |
| CHA<sub>2</sub>DS<sub>2</sub>-VASc | Algorithm | Expert B | 0.80 (0.68-0.92) |
| CHA<sub>2</sub>DS<sub>2</sub>-VASc | Expert A | Expert B | 0.85 (0.73-0.97) |
| HAS-BLED | Algorithm | Expert A | 0.54 (0.36-0.72) |
| HAS-BLED | Algorithm | Expert B | 0.53 (0.34-0.72) |
| HAS-BLED | Expert A | Expert B | 0.74 (0.51-0.97) |

*Raters 1 and 2 are two independent clinician raters, Algorithm is the automatic scoring pipeline developed in this paper.*

### Temporal Trends in Antithrombotic drug prescription

Prior to 2013, OAC use varied between 40-45% (mean 43.4%) with no strong trend (linear regression R^2^=0.08, slope = +0.2% per quarter, Fig 2a,b). From 2013 onwards the average OAC rate remained above 47% and there was a gradual increase in OAC use such that at the end of the study period 68.4% of patients were taking an OAC (linear regression R^2^=0.77, slope = +1.2% per quarter). This increase in OAC rate is particularly pronounced from 2016 onwards (linear regression R^2^=0.86, slope = +2.7% per quarter). Conversely, the proportion of patients taking an AP drug alone declined significantly from 48.9% at the start to 14.5% at the end of the study, with a consistent linear decrease over the period (linear regression R^2^=0.94, slope = −1.24% per quarter).

**Fig 2.**
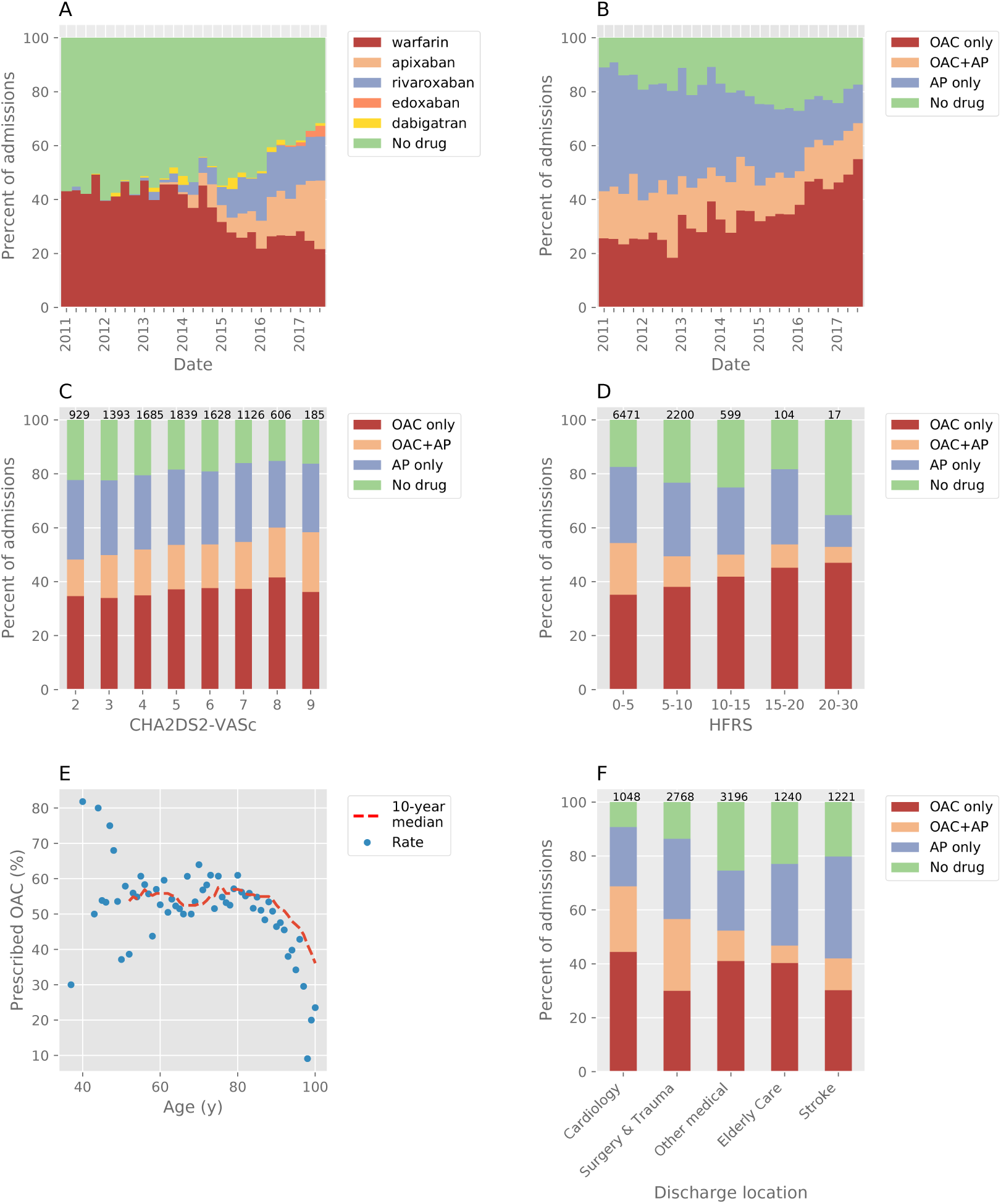
Antithrombotic drug prescribing patterns in the AF cohort patients with CHA_2_DS_2_-VASc ≥ 2. A,B) Prescribing rates for all admissions during the study period. A) OAC choice vs. no OAC. B) Prescribing of OAC and/or AP vs. neither. C) Prescribing rates stratified by CHA_2_DS_2_-VASc for all patients. D) Prescribing rates grouped by HFRS as defined by Gilbert et al. Due to low numbers of patients with score > 20 the final (highest) bin is wider than the others. E) Prescribing rate vs. age at discharge. Points are the mean prescribing rate per year for all ages with ≥ 10 patients, a 10-year moving median (trend) is shown as a dashed red line. F) prescribing rates in patients grouped by discharging specialty. In C, D, F the number above each bar indicates the number of patients. AP = antiplatelet, HFRS = hospital frailty risk score, OAC = oral anticoagulant.

At the start of the study warfarin was the only widely available OAC. In 2012 NICE endorsed the use of the first 2 DOACs (Dabigatran and Rivaroxaban) and the prescription of both drugs increased from the end of 2012, at a similar time to when overall OAC use began to rise. From then on there was a gradual increase in the use of DOACs at the expense of warfarin, such that at the end of the study period in 2017 warfarin only contributed a third of all OAC prescriptions.

For newly diagnosed AF (n=4986) Antithrombotic drug trends closely mirrored those found in the overall AF cohort (Fig 3).

**Fig 3.**
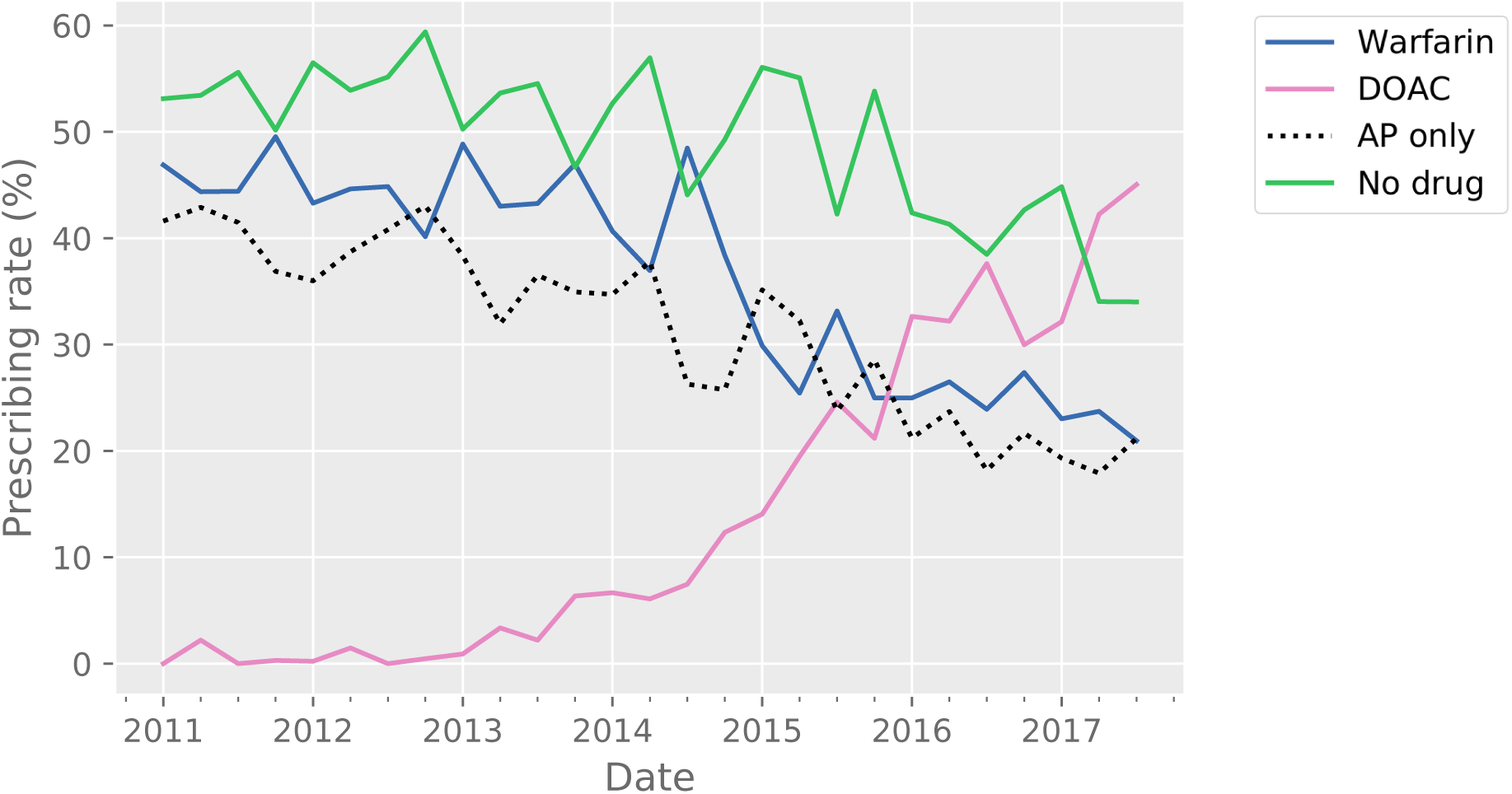
Prescribing trends for new AF cases over the study period. The solid blue line represents warfarin, the solid pink line represents DOAC, the dashed black line represents AP prescription without any OAC, the solid green line represents the no drug group. Total N = 4986. AP = antiplatelet, DOAC = direct oral anticoagulant, OAC = oral anticoagulant.

### Clinical Factors associated with Antithrombotic drug prescription

There was gradual increase in rates of OAC use with a higher CHA_2_DS_2_-VASc score (+1.6% per point, linear regression R^2^ = 0.93, p < 0.001) (Fig 2c). Conversely OAC prescription decreased with older age in patients ≥80 years (Fig 2e).

In multivariate analysis (Table 4) clinical variables associated with a higher rate of OAC use (vs. no OAC) included heart failure, diabetes and stroke. Factors negatively associated with OAC use included a history of vascular disease, abnormal liver function and history of bleeding. Older patients receiving OAC were more likely to be on warfarin vs. DOACs. Higher rates of AP drug use alone (vs. OAC) were associated with the presence of vascular disease, whereas heart failure, and diabetes were associated with lower rates.

**Table 4.** Univariate and multivariate logistic regression for factors associated with antithrombotic drug prescribing at most recent discharge for patients with CHA2DS2-VASc ≥ 2.

| Group | Factor | Any OAC vs no OAC |  |  |  | DOAC vs Warfarin |  |  |  | AP-only vs OAC-only |  |  |  |
| --- | --- | --- | --- | --- | --- | --- | --- | --- | --- | --- | --- | --- | --- |
|  |  | Univariate |  | Multivariate |  | Univariate |  | Multivariate |  | Univariate |  | Multivariate |  |
|  |  | OR (95%CI) | P-value | OR (95%CI) | P-value | OR (95%CI) | P-value | OR (95%CI) | P-value | OR (95%CI) | P-value | OR (95%CI) | P-value |
| *Other clinical variables* | Age (per 20 years) | 0.9<br>(0.9-1.0) | 0.0<br>39 | 0.9<br>(0.8-0.9) | <0.<br>001 | 1.3<br>(1.2-1.4) | <0.<br>001 | 0.8<br>(0.8-0.9) | <0.<br>001 | 1.4<br>(1.2-1.5) | <0.<br>001 | 1.0<br>(1.0-1.1) | 0.0<br>80 |
|  | LOS (per 14 days) | 0.9<br>(0.9-1.0) | <0.<br>001 | 1.0<br>(0.9-1.0) | 0.0<br>16 | 1.1<br>(1.1-1.2) | <0.<br>001 | 1.1<br>(1.0-1.1) | 0.0<br>25 | 1.1<br>(1.0-1.1) | 0.0<br>06 | 1.0<br>(1.0-1.1) | 0.0<br>73 |
|  | Visits (per 10) | 1.1<br>(1.0-1.1) | <0.<br>001 | 1.1<br>(1.1-1.1) | <0.<br>001 | 1.1<br>(1.1-1.1) | <0.<br>001 | 1.0<br>(1.0-1.1) | 0.4<br>46 | 0.9<br>(0.9-1.0) | <0.<br>001 | 0.9<br>(0.9-1.0) | <0.<br>001 |
| *CHA2DS2-VASc factors* | Congestive heart failure | 1.7<br>(1.6-1.8) | <0.<br>001 | 1.7<br>(1.5-1.8) | <0.<br>001 | 1.0<br>(0.9-1.1) | 0.8<br>99 |  |  | 0.7<br>(0.6-0.8) | <0.<br>001 | 0.7<br>(0.6-0.8) | <0.<br>001 |
|  | Diabetes mellitus | 1.4<br>(1.3-1.5) | <0.<br>001 | 1.2<br>(1.1-1.3) | <0.<br>001 | 1.0<br>(0.9-1.1) | 0.9<br>73 |  |  | 0.8<br>(0.7-0.9) | <0.<br>001 | 0.9<br>(0.8-1.0) | 0.0<br>33 |
|  | Female | 1.0<br>(0.9-1.0) | 0.3<br>27 |  |  | 1.2<br>(1.1-1.4) | 0.0<br>02 | 1.1<br>(1.0-1.2) | 0.1<br>69 | 1.1<br>(1.0-1.2) | 0.2<br>21 |  |  |
|  | Hypertension | 1.1<br>(1.0-1.2) | 0.0<br>42 | 1.1<br>(1.0-1.2) | 0.1<br>37 | 1.4<br>(1.2-1.5) | <0.<br>001 | 1.1<br>(0.9-1.2) | 0.2<br>54 | 1.1<br>(1.0-1.2) | 0.0<br>89 |  |  |
|  | Stroke | 1.1<br>(1.0-1.2) | 0.0<br>20 | 1.3<br>(1.1-1.4) | <0.<br>001 | 1.4<br>(1.3-1.6) | <0.<br>001 | 1.0<br>(0.9-1.2) | 0.6<br>69 | 1.0<br>(0.9-1.1) | 0.5<br>51 |  |  |
|  | Vascular disease | 1.1<br>(1.0-1.2) | 0.0<br>18 | 0.9<br>(0.8-0.9) | 0.0<br>03 | 1.0<br>(0.9-1.1) | 0.6<br>85 |  |  | 1.3<br>(1.1-1.5) | <0.<br>001 | 1.6<br>(1.4-1.9) | <0.<br>001 |
| *HAS-BLED factors* | Abnormal liver function | 0.7<br>(0.6-0.9) | <0.<br>001 | 0.7<br>(0.5-0.8) | <0.<br>001 | 1.0<br>(0.8-1.3) | 0.9<br>52 |  |  | 1.1<br>(0.8-1.4) | 0.5<br>59 |  |  |
|  | Abnormal renal function | 1.1<br>(1.0-1.2) | 0.1<br>36 |  |  | 1.3<br>(1.1-1.5) | 0.0<br>02 | 1.0<br>(0.8-1.1) | 0.5<br>94 | 0.9<br>(0.8-1.0) | 0.1<br>17 |  |  |
|  | Bleeding | 0.6<br>(0.5-0.7) | <0.<br>001 | 0.6<br>(0.5-0.6) | <0.<br>001 | 1.3<br>(1.1-1.5) | 0.0<br>14 | 0.9<br>(0.8-1.2) | 0.6<br>20 | 1.2<br>(1.0-1.4) | 0.0<br>81 |  |  |
| *Frailty* | HFRS (per 10 points) | 0.8<br>(0.7-0.9) | <0.<br>001 | 0.7<br>(0.6-0.8) | <0.<br>001 | 2.6<br>(2.2-3.0) | <0.<br>001 | 2.1<br>(1.8-2.6) | <0.<br>001 | 1.2<br>(1.0-1.4) | 0.0<br>15 | 1.2<br>(1.0-1.4) | 0.0<br>41 |
| *Discharge Location* | Stroke | 0.6<br>(0.6-0.7) | <0.<br>001 | (reference) |  | 1.4<br>(1.1-1.6) | <0.<br>001 | (reference) |  | 2.2<br>(1.9-2.5) | <0.<br>001 | (reference) |  |
|  | Cardiology | 2.2<br>(2.0-2.5) | <0.<br>001 | 2.6<br>(2.2-3.0) | <0.<br>001 | 0.7<br>(0.6-0.8) | <0.<br>001 | 0.5<br>(0.4-0.7) | <0.<br>001 | 0.3<br>(0.3-0.4) | <0.<br>001 | 0.2<br>(0.2-0.3) | <0.<br>001 |
|  | Elderly Care | 0.8<br>(0.7-0.9) | <0.<br>001 | 1.2<br>(1.0-1.4) | 0.0<br>36 | 1.9<br>(1.6-2.2) | <0.<br>001 | 0.8<br>(0.7-1.1) | 0.2<br>34 | 1.2<br>(1.0-1.4) | 0.0<br>13 | 0.6<br>(0.5-0.7) | <0.<br>001 |
|  | Other medical specialties | 0.8<br>(0.8-0.9) | <0.<br>001 | 1.2<br>(1.0-1.4) | 0.0<br>13 | 1.2<br>(1.0-1.3) | 0.0<br>23 | 0.7<br>(0.5-0.8) | <0.<br>001 | 1.0<br>(0.9-1.1) | 0.9<br>05 | 0.6<br>(0.5-0.7) | <0.<br>001 |
|  | Surgery & Trauma | 1.2<br>(1.1-1.3) | <0.<br>001 | 1.6<br>(1.4-1.8) | <0.<br>001 | 0.7<br>(0.6-0.8) | <0.<br>001 | 0.5<br>(0.4-0.6) | <0.<br>001 | 0.8<br>(0.7-1.0) | 0.0<br>13 | 0.5<br>(0.4-0.5) | <0.<br>001 |

All factors significant at $p < 0.05$ level in univariate analysis were included in the multivariate model. HFRS = hospital frailty risk score, LOS = length of stay

### Hospital Frailty Risk Score (HFRS) and antithrombotic prescription

As HFRS increased, OAC use did not significantly change but there was a clear decrease in AP drug use either alone or with an OAC (−8.3% per group, linear regression R^2^ = 0.85, p < 0.01, Fig 2d). However in multivariate analysis increasing HFRS was strongly negatively associated with OAC use, positively associated with DOAC use and positively associated with AP drug use only.

### Relationship Between Discharging Specialty and OAC use

We found a large variation in OAC prescribing rates between different specialities (Fig 2f). The highest rate of OAC use was in patients discharged from cardiology (68.8%, n=1048), with lower rates of OAC use in patients discharged under a surgical team (56.6%, n=2768), a medical specialty (52.3%, n=3196), elderly care (46.8%, n=1249) and the stroke unit (42.0%, n=1222). The relationship between discharge location and antithrombotic drug use remained significant after correction for a range of clinical variables, age and HFRS (Table 4).

### Medication switching in AF patients

We identified a group of 1708 patients (CHA_2_DS_2_-VASc ≥ 2) with 2 or more admissions at least 12 months apart. Of these 895 (52.4%) changed their antithrombotic medication status (Fig 4a). Overall there was an increase in OAC use from 985 to 1069 patients (+8.5%) and a net movement of patients to DOACs from warfarin and AP drugs. These findings were more marked when only patients whose admissions straddled the 2014 NICE guidelines update were included (1096 patients; Fig 4b).

**Fig 4.**
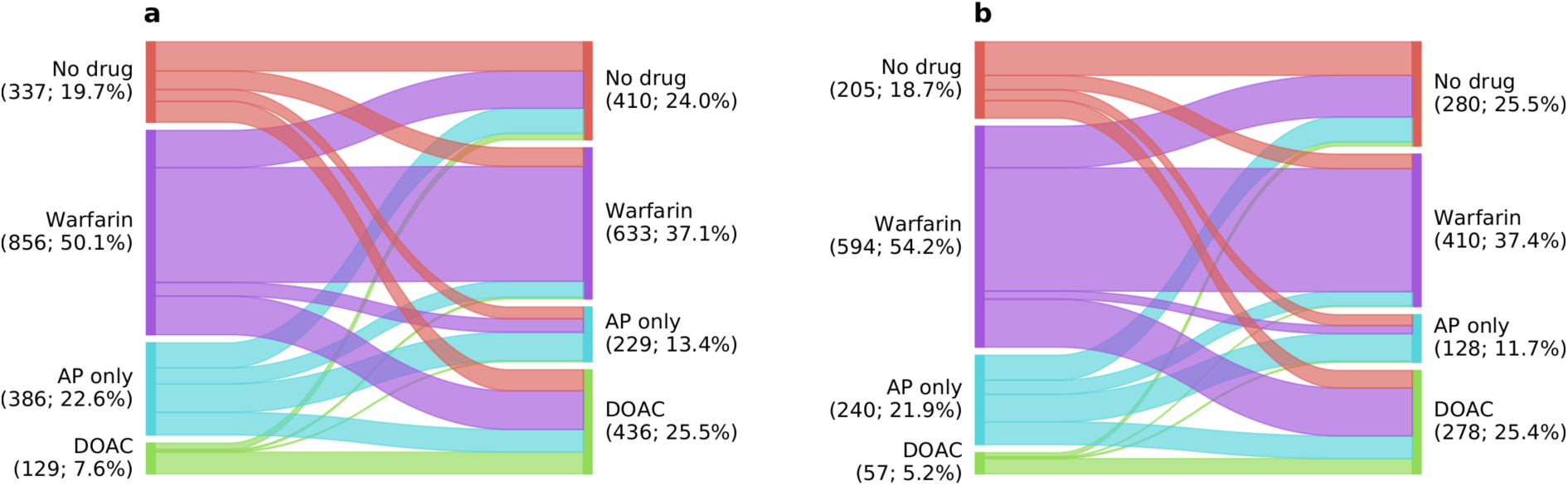
Medication switching in patients with CHA_2_DS_2_-VASc ≥ 2 at last visit. a) all visits at least 12 months apart and b) last visit before vs last visit after the 2014 NICE guideline update (b is a subset of a). Line width indicates overall proportion.

## Discussion

We have developed a pipeline to calculate clinical risk scores from free-text using NLP. Using this pipeline, we were able to estimate CHA_2_DS_2_-VASc and HAS-BLED risk scores from free-text EHR data that are in line with those calculated manually and could scale up to analyse data on over 10,000 AF patients managed at a multi-site large UK NHS Trust.

We were able to replicate the changes in antithrombotic drug practices observed over the last 7 years in previous registry-based observational studies. First, there has been a substantial increase in the proportion of AF patients at high risk of stroke (CHA_2_DS_2_-VASc ≥ 2) prescribed an OAC, with OAC use rising from 42% in 2011 to 62% in 2017. Second, there has been a reduction in the use of warfarin and an increase in DOAC prescription, such that in 2017 more patients were discharged on a DOAC than warfarin. Third, the use of AP drugs alone to prevent stroke has dropped significantly, from 40% in 2011 to 10% in 2017.

### Semantic NLP analysis of routinely-generated clinical data

Clinical applications of NLP are an active research area. A recent systematic review identified 71 NLP applications for clinical text, 12 of which are open-source.[22] We took different approaches to NLP for the two major components of our study: extracting medication from discharge summaries and detecting clinical concepts in text (to derive risk scores). For medications, we use a series of regular expression rules tuned to the specific prescription text used in this study with high precision but less generalizability. For risk scoring, we built a concept mapping pipeline on top of an open-source clinical NLP tool SemEHR[13], which can detect far more concepts than it is feasible to manually code rules for, but with the trade-off that it is not specifically designed for any particular disease concepts.

### Use of EHR data for retrospective and prospective applications in cardiology

EHRs have been increasingly used to support observational studies. However, typically this involves the transcription of clinical data from EHRs into a registry-specific electronic case report form, an approach with many of the limitations inherent of a classical observational study. The development and maintenance of case registries is time-consuming, and the scope of the research questions that can be answered are limited to the dataset defined *a priori*. By using a domain-agnostic concept mapping pipeline (SemEHR) on unstructured text, our study was able to test both conventional risk scores (CHA_2_DS_2_-VASc) and a novel risk score (HFRS).

Ours is not the first study to utilize unstructured EHR data in AF research.[15–17,23] Our study builds on this previous work through the use of text data with an NLP pipeline, the calculation of additional risk scores and an analysis of prescribing patterns. Whilst we evaluate our pipeline in the context of AF, our aim is to provide an open tool for clinical risk scoring calculations in general.

### Trends in Antithrombotic drug use

Large retrospective population-based studies have established a clear trend of increased OAC prescribing in AF patients, driven by uptake of DOACs.[6,7] Our ability to reproduce these findings by applying NLP to unstructured EHR data strongly supports the validity of the NLP pipeline. In our analysis, OAC prescription was independently associated with risk factors for stroke and bleeding, consistent with the findings of other studies.

Despite a significant increase in OAC use during our study period, ∼35% of patients at high risk of stroke were still not prescribed an OAC indicating there are some remaining barriers to OAC use. In our data, a documented bleeding problem (present in 14% of the cohort and associated with 40% reduction in OAC use) and increasing frailty (Table 4) were independent predictors of OAC underuse, suggesting that perceived risk of bleeding and risk of harm due to OAC continues, particularly in elderly patients, to have a strong influence on the antithrombotic drug decision-making process.[24–26]

HFRS proposed by Gilbert *et al*. [20] is a high-dimensional frailty score calculated from ICD-10 diagnostic codes. When we evaluated antithrombotic drug prescription using HFRS as a continuous variable and adjusting for other clinical variables and discharging specialty, there was a significant relationship between HFRS and antithrombotic drug use (Table 4). Patients with a higher HFRS were less likely to take an OAC, more likely to take a DOAC (vs. warfarin) if they were on an OAC, and more likely to take an AP drug alone versus an OAC. This suggests there is an underlying high-dimensional frailty characteristic influencing clinician decision-making despite not being explicitly calculated.

The highest OAC prescription rates were in patients discharged from a cardiology ward (n=1048, 69%), whereas OAC use was significantly lower in patients discharged from an elderly care ward (n=1240, 47%) and other medical specialties (n=3196, 52%). Although in part this may reflect the differing case mix of specialty patient populations, given the magnitude of the differences seen even with multivariate correction of clinical variables (including stroke and bleed risk factors and frailty risk score), it is likely that some of our findings are due to specialty-specific behaviours in relation to AF and bleeding risk. This suggests efforts to continue to increase OAC prescribing rates beyond current may be most effective if targeted by clinical specialty.

### Limitations

One of the major limitations of an EHR- and NLP-based approach, as used in our analysis, is data accuracy. We manually validated the major variables in our analysis but the accuracy of our NLP algorithm deserves closer scrutiny as there is a risk of causing a significant degradation in data accuracy. Whilst the agreement between our algorithm and clinical experts was high for CHA2DS2-VASc and fair for HAS-BLED, in all comparisons the agreement between experts was higher. This gap represents room for improvement in the algorithm primarily due to difficulty detecting some risk factors.

Retrospective assessment of the data source of many of the variables in the HAS-BLED score is challenging irrespective of the approach used, with a previous study finding that inter-rater reliability between human observers for some HAS-BLED components is low.[15] This disagreement at the level of the data source is commonly described even with curated registry data.[27] This limitation particularly affected the “uncontrolled hypertension” and “labile INR” features of the HAS-BLED score, neither of which is reliably recorded or detected. This leaves some comorbidity associated with bleeding risk unaccounted for in our multivariate analysis.

Unlike the use of registry data, routine EHR data may not capture all necessary clinical information on all patients, as this is a secondary use of the record. It is therefore possible that we have missed important co-morbidities in some of the patients. This may have led to an overall underestimation of co-morbidities in our patient population, as well as undermined some of our analyses relating clinical variables to anti-thrombotic drug use.

The NLP algorithm was tested on data from one multi-site organization using three different EHR systems over a 6-year period. While this may show a degree of generalizability, further validation on data from other EHR systems in other organizations will be needed.

We used data from inpatient admissions as these more accurately record data on drug prescriptions. As a result our patient population has the potential to be older and frailer, with more comorbidity, than typical community AF cohorts. Although our population had similar baseline characteristics to the populations in previous studies[28,29], not all co-morbidities may be captured. This is a limitation is inherent in the design of all studies using routinely generated non-curated data.

Our study did not attempt to distinguish between the different temporal patterns of atrial fibrillation (permanent, persistent, paroxysmal). This is because these temporal patterns are frequently not used in free text or used ambiguously (e.g. ‘PAF’ could mean any of the terms). Nonetheless, national and international guidelines on anticoagulation for AF do not have different anticoagulation recommendations for different temporal patterns.

Finally, our data is observational. Therefore, although we have demonstrated associations between changes in antithrombotic drug use and a range of clinical variables, it is not possible to conclude a causal link.

## Conclusion

We present a novel open-source methodology for an automated pipeline to calculate risk scores from NLP and track prescribing patterns, incorporating future disease entities, risk profiles and ontologies. We have used this methodology to demonstrate significant changes in antithrombotic practice in AF since the introduction of DOACs, in a large NHS Trust. The tools used in this study are open-source and transparent (CogStack[12], SemEHR[14] and our pipeline) allowing any other organization to validate on their own cohorts and optimize local population health at low cost. This highlights the power of semantic NLP processing tools for a disease-specific domain, but is generalizable to a variety of other diseases and use-cases, and highlights the growing impact of health informatics in healthcare.[30]

## Data Availability

Source text from patient records used in the study will not be available due to inability to fully anonymise up to the Information Commissioner Office (ICO) standards and would be likely to contain strong identifiers (e.g. names, postcodes) and highly sensitive data (e.g. diagnoses). A subset of the dataset limited to anonymisable information (e.g. only UMLS codes and demographics) is available on request to researchers with suitable training in information governance and human confidentiality protocols subject to approval by the King’s College Hospital Information Governance committee; applications for research access should be sent to. This dataset cannot be released publicly due to the risk of re-identification of such granular individual-level data, as determined by the King’s College Hospital Caldicott Guardian. All code for calculating risk scores is open-source in GitHub at "https://github.com/CogStack/risk-score-builder".

## Abbreviations

AF: atrial fibrillation
AP: antiplatelet
DOAC: direct oral anticoagulant
EHR: electronic health record
NLP: natural language processing
OAC: oral anticoagulant

## Acknowledgements

DMB is funded by a UKRI Innovation Fellowship as part of Health Data Research UK MR/S00310X/1 (https://www.hdruk.ac.uk). HW is funded by a UKRI Rutherford Fellowship as part of Health Data Research UK MR/S004149/1. RB is funded in part by grant MR/R016372/1 for the King’s College London MRC Skills Development Fellowship programme funded by the UK Medical Research Council (MRC, https://mrc.ukri.org) and by grant IS-BRC-1215-20018 for the National Institute for Health Research (NIHR, https://www.nihr.ac.uk) Biomedical Research Centre at South London and Maudsley NHS Foundation Trust and King’s College London. AMS is supported by the British Heart Foundation (https://www.bhf.org.uk). NIHR Biomedical Research Centre funding to SLAM/KCL and to GSTT/KCL in partnership with KCL. RJBD is supported by: 1. Health Data Research UK, which is funded by the UK Medical Research Council, Engineering and Physical Sciences Research Council, Economic and Social Research Council, Department of Health and Social Care (England), Chief Scientist Office of the Scottish Government Health and Social Care Directorates, Health and Social Care Research and Development Division (Welsh Government), Public Health Agency (Northern Ireland), British Heart Foundation and Wellcome Trust. 2. The BigData@Heart Consortium, funded by the Innovative Medicines Initiative-2 Joint Undertaking under grant agreement No. 116074. This Joint Undertaking receives support from the European Union’s Horizon 2020 research and innovation programme and EFPIA; it is chaired, by DE Grobbee and SD Anker, partnering with 20 academic and industry partners and ESC. 3. The National Institute for Health Research University College London Hospitals Biomedical Research Centre. 4. National Institute for Health Research (NIHR) Biomedical Research Centre at South London and Maudsley NHS Foundation Trust and King’s College London.

This paper represents independent research part funded by the National Institute for Health Research (NIHR) Biomedical Research Centre at South London and Maudsley NHS Foundation Trust and King’s College London. The views expressed are those of the author(s) and not necessarily those of the NHS, the NIHR or the Department of Health and Social Care. The funders had no role in study design, data collection and analysis, decision to publish, or preparation of the manuscript.

## Competing Interests

I have read the journal’s policy and the authors of this manuscript have the following competing interests: Dr. Teo reports non-financial support from Bayer, grants from Bristol-Meyers-Squibb, outside the submitted work; Dr. scott reports personal fees from Bayer, outside the submitted work. All other authors declare that no competing interests exist. This does not alter our adherence to PLOS ONE policies on sharing data and materials.

## Supporting information

**S1 Table. Definition of HAS-BLED and CHA2DS2-VASc as used in this study**. Age and gender are included directly from electronic health record data. The agreed terms under “include” and “exclude” headings were used by clinical experts to calculate each score manually. The lists of UMLS concepts for each component were derived automatically and used by the NLP scoring algorithm.

**S2 Table. Performance of the NLP pipeline for each component of CHA**_**2**_**DS**_**2**_**-VASc and HAS-BLED**. Cases were considered positive if at least one manual rater marked as positive. The agreement between the two manual raters is shown as “agreement between raters”.

**S3 Table. Total score and component score for CHA**_**2**_**DS**_**2**_**-VASc and HAS-BLED**. Each row represents a single patient identified only by row number (“Patient” column).

## References

1. Yiin GSC, Howard DPJ, Paul NLM, Li L, Mehta Z, Rothwell PM, et al. Recent time trends in incidence, outcome and premorbid treatment of atrial fibrillation-related stroke and other embolic vascular events: a population-based study. J Neurol Neurosurg Psychiatry. 2015/10/20. 2017;88: 12–18. doi:10.1136/jnnp-2015-311947

2. NICE. Atrial fibrillation: management (Aug 2014 update) [Internet]. 2014.

3. Lip GYH, Nieuwlaat R, Pisters R, Lane DA, Crijns HJGM. Refining Clinical Risk Stratification for Predicting Stroke and Thromboembolism in Atrial Fibrillation Using a Novel Risk Factor-Based Approach: The Euro Heart Survey on Atrial Fibrillation. Chest. 2010;137: 263–272. doi:10.1378/chest.09-1584

4. Pisters R, Lane DA, Nieuwlaat R, de Vos CB, Crijns HJGM, Lip GYH. A Novel User-Friendly Score (HAS-BLED) To Assess 1-Year Risk of Major Bleeding in Patients With Atrial Fibrillation: The Euro Heart Survey. Chest. 2010;138: 1093–1100. doi:10.1378/chest.10-0134

5. Cowan C, Healicon R, Robson I, Long WR, Barrett J, Fay M, et al. The use of anticoagulants in the management of atrial fibrillation among general practices in England. Heart. 2013;99: 1166–1172. doi:10.1136/heartjnl-2012-303472

6. Campbell Cowan J, Wu J, Hall M, Orlowski A, West RM, Gale CP. A 10 year study of hospitalized atrial fibrillation-related stroke in England and its association with uptake of oral anticoagulation. Eur Heart J. 2018; doi:10.1093/eurheartj/ehy411

7. Lacoin L, Lumley M, Ridha E, Pereira M, McDonald L, Ramagopalan S, et al. Evolving landscape of stroke prevention in atrial fibrillation within the UK between 2012 and 2016: a cross-sectional analysis study using CPRD. BMJ Open. 2017;7: e015363. doi:10.1136/bmjopen-2016-015363

8. Holt TA, Hunter TD, Gunnarsson C, Khan N, Cload P, Lip GYH. Risk of stroke and oral anticoagulant use in atrial fibrillation: A cross-sectional survey. Br J Gen Pract. 2012; doi:10.3399/bjgp12X656856

9. Hemingway H, Asselbergs FW, Danesh J, Dobson R, Maniadakis N, Maggioni A, et al. Big data from electronic health records for early and late translational cardiovascular research: challenges and potential. Eur Heart J. 2017;39: 1481–1495. doi:10.1093/eurheartj/ehx487

10. Kharrazi H, Anzaldi LJ, Hernandez L, Davison A, Boyd CM, Leff B, et al. The Value of Unstructured Electronic Health Record Data in Geriatric Syndrome Case Identification. J Am Geriatr Soc. 2018;66: 1499–1507. doi:10.1111/jgs.15411

11. Jackson R, Kartoglu I, Stringer C, Gorrell G, Roberts A, Song X, et al. CogStack - Experiences of deploying integrated information retrieval and extraction services in a large National Health Service Foundation Trust hospital. BMC Med Inform Decis Mak. 2018; doi:10.1186/s12911-018-0623-9

12. CogStack. CogStack Pipeline [Internet]. 2019. Available: https://github.com/CogStack/CogStack-Pipeline

13. Wu H, Toti G, Morley KI, Ibrahim ZM, Folarin A, Jackson R, et al. SemEHR: A general-purpose semantic search system to surface semantic data from clinical notes for tailored care, trial recruitment, and clinical research. J Am Med Informatics Assoc. 2018; doi:10.1093/JAMIA/OCX160

14. Wu H. CogStack-SemEHR [Internet]. p. 2019.

15. Wang S V, Rogers JR, Jin Y, Fischer MA, Bates DW. Use of electronic healthcare records to identify complex patients with atrial fibrillation for targeted intervention. J Am Med Informatics Assoc. 2016;24: 339–344. doi:10.1093/jamia/ocw082

16. Grouin C, Deléger L, Rosier A, Temal L, Dameron O, Van Hille P, et al. Automatic computation of CHA2DS2-VASc score: information extraction from clinical texts for thromboembolism risk assessment. AMIA. Annu Symp proceedings AMIA Symp. 2011;

17. Rosier A, Mabo P, Temal L, Van Hille P, Dameron O, Deléger L, et al. Personalized and automated remote monitoring of atrial fibrillation. Europace. 2016; doi:10.1093/europace/euv234

18. U.S. National Library of Medicine. Unified Medical Language System (UMLS) [Internet]. Available: https://www.nlm.nih.gov/research/umls/

19. Köhler S, Carmody L, Vasilevsky N, Jacobsen JOB, Danis D, Gourdine JP, et al. Expansion of the Human Phenotype Ontology (HPO) knowledge base and resources. Nucleic Acids Res. 2019; doi:10.1093/nar/gky1105

20. Gilbert T, Neuburger J, Kraindler J, Keeble E, Smith P, Ariti C, et al. Development and validation of a Hospital Frailty Risk Score focusing on older people in acute care settings using electronic hospital records: an observational study. Lancet. 2018; doi:10.1016/S0140-6736(18)30668-8

21. Whetzel PL, Noy NF, Shah NH, Alexander PR, Nyulas C, Tudorache T, et al. BioPortal: Enhanced functionality via new Web services from the National Center for Biomedical Ontology to access and use ontologies in software applications. Nucleic Acids Res. 2011; doi:10.1093/nar/gkr469

22. Kreimeyer K, Foster M, Pandey A, Arya N, Halford G, Jones SF, et al. Natural language processing systems for capturing and standardizing unstructured clinical information: A systematic review. Journal of Biomedical Informatics. 2017. doi:10.1016/j.jbi.2017.07.012

23. Piazza G, Hurwitz S, Galvin CE, Harrigan L, Baklla S, Hohlfelder B, et al. Alert-based computerized decision support for high-risk hospitalized patients with atrial fibrillation not prescribed anticoagulation: a randomized, controlled trial (AF-ALERT). Eur Heart J. 2019; doi:10.1093/eurheartj/ehz385

24. Bahri O, Roca F, Lechani T, Druesne L, Jouanny P, Serot J-M, et al. Underuse of Oral Anticoagulation for Individuals with Atrial Fibrillation in a Nursing Home Setting in France: Comparisons of Resident Characteristics and Physician Attitude. J Am Geriatr Soc. 2015;63: 71–76. doi:10.1111/jgs.13200

25. Lefebvre M-CD, St-Onge M, Glazer-Cavanagh M, Bell L, Kha Nguyen JN, Viet-Quoc Nguyen P, et al. The Effect of Bleeding Risk and Frailty Status on Anticoagulation Patterns in Octogenarians With Atrial Fibrillation: The FRAIL-AF Study. Can J Cardiol. 2016;32: 169–176. doi:10.1016/j.cjca.2015.05.012

26. Pilotto A, Gallina P, Copetti M, Pilotto A, Marcato F, Mello AM, et al. Warfarin Treatment and All-Cause Mortality in Community-Dwelling Older Adults with Atrial Fibrillation: A Retrospective Observational Study. J Am Geriatr Soc. 2016/06/13. 2016;64: 1416–1424. doi:10.1111/jgs.14221

27. Faxon DP, Burgess A. Cardiovascular Registries: Too Much of Good Thing? Circulation. Cardiovascular interventions. United States; 2016. p. e003866. doi:10.1161/CIRCINTERVENTIONS.116.003866

28. Marzec LN, Wang J, Shah ND, Chan PS, Ting HH, Gosch KL, et al. Influence of Direct Oral Anticoagulants on Rates of Oral Anticoagulation for Atrial Fibrillation. J Am Coll Cardiol. 2017;69: 2475–2484. doi: https://doi.org/10.1016/j.jacc.2017.03.540

29. Fosbol EL, Holmes DN, Piccini JP, Thomas L, Reiffel JA, Mills RM, et al. Provider specialty and atrial fibrillation treatment strategies in United States community practice: findings from the ORBIT-AF registry. J Am Heart Assoc. 2013;2: e000110–e000110. doi:10.1161/JAHA.113.000110

30. Topol EJ. High-performance medicine: the convergence of human and artificial intelligence. Nat Med. 2019;25: 44–56. doi:10.1038/s41591-018-0300-7

